# Vision Impairment prediction for patients diagnosed with Multiple Sclerosis: Cosmos based machine learning model training and evaluation

**DOI:** 10.1101/2023.11.10.23298366

**Authors:** Brandon Buxton, Amr Hassan, Nevin Shalaby, John Lindsey, John Lincoln, Elmer Bernstam, Wagida Anwar, Degui Zhi, Laila Rasmy

## Abstract

**Objectives:** Multiple sclerosis (MS) is a complex autoimmune neurological disorder that frequently impacts vision. One of the most frequent initial presentations of MS is acute vision loss due to optic neuritis, an acute disorder caused by MS involvement with the optic nerve. While vision impairment is often the first sign of MS, it can occur or recur at any time during the patient’s course. In this study, we aim to develop and evaluate machine learning models to predict vision impairment in patients with MS, both at the time of first MS diagnosis and throughout their course of care. Early awareness and intervention in patients likely to have vision loss can help preserve patient quality of life.

**Materials and Methods:** Using the Epic Cosmos de-identified electronic health record (EHR) dataset, we queried 213+ million patients to extract our MS cohort. Cases were defined as MS patients with vision impairment or optic neuritis (VI) following their first MS diagnosis, while controls were MS patients without VI. We trained logistic regression (LR), light gradient boosting machine (LGBM), and recurrent neural network (RNN) models to predict future VI in MS patients. The models were evaluated for two distinct clinical tasks: prediction of VI at the time of the first MS diagnosis and prediction of VI at the most recent visit. Similarly, we trained the models on different segments of the patient trajectory including up until the first MS diagnosis (MS-First Diagnosis), or until the most recent visit before developing the outcome (MS-Progress) as well as the combination of both (MS-General). Finally, we trained a survival model with the goal of predicting patient likelihood of vision loss over time. We compared the models’ performance using AUROC, AUPRC, and Brier scores.

**Results:** We extracted a cohort of 377,097 patients with MS, including 42,281 VI cases. Our trained models achieved ∼80% AUROC, with RNN-based models outperforming LGBM and LR (79.6% vs 72.8% and 68.6%, respectively) when considering the full patient trajectory. The MS-General RNN model had the highest AUROC (64.4%) for predicting VI at the first MS diagnosis. The MS-Progress survival model achieved a 75% concordance index on the full trajectory, while the more clinically relevant MS-First Diagnosis model achieved 63.1% at initial diagnosis.

**Discussion and Conclusion:** The MS-Progress and MS-General RNN models performed best in both prediction scenarios. While MS-General achieved the best performance at the time of first MS diagnosis with around 1% AUROC increase compared to the MS-First Diagnosis model, it showed around 1% AUROC decrease on the MS progress scenario. All RNN survival models performed the best when they were trained on data corresponding to the evaluation use-case scenarios. RNN based models showed promising performance that demonstrates that they can be useful clinical tools to predict risk of future VI events in patients with MS. Further development of these models will focus on expanding to predict other comorbidities associated with MS relapse or progression.

## INTRODUCTION

Multiple sclerosis (MS) is a progressive neurologic disease caused by demyelination of central nervous system (CNS) axons, typically first presenting in patients under 50 years old.^1^ Patients with MS may experience a heterogenous spectrum of neurologic symptoms, including sensory loss, paresthesias, motor disturbances, ataxia, difficulties with balance, and incontinence.^2^ Frequently, the initial presentation of MS features vision disturbances ranging from an isolated decrease in visual acuity to optic neuritis (ON), a condition typically featuring worsening vision loss over 7-10 days, color desaturation, and pain with ocular movement.^3^ The seminal 1988-1991 Optic Neuritis Treatment Trial (ONTT) demonstrated that previously undiagnosed patients, onset of ON and presence of at least one demyelinated lesion on brain MRI resulted in 72% risk of developing MS over the following 15 years.^4^ Isolated ON may be the initial presentation of MS in up to 20% of patients, with overall ON prevalence of ∼50% throughout the course of the disease.^5^ Despite the strong association between ON and MS, optic nerve lesions seen on neuroimaging are not included in the latest MS diagnostic criteria.^6^

While the relationship between vision impairment and MS diagnosis has been well documented, relatively little has been established to predict the risk of vision impairment in patients with MS who do not present with vision loss prior to or concurrently with MS diagnosis. These causes can be both complications of ON and ocular disorders independent of ON status.^7^ Retinal periphlebitis, macular thinning, uveitis, and multiple other conditions commonly found in the general population (glaucoma, cataracts, etc.) are known to be prevalent in patients with MS.^8^ These can develop independently of optic neuritis and cause continued vision degradation without acute ON. Efferent manifestations of MS can also affect visuomotor pathways, leading to abnormal eye movements, diplopia, and blurred vision. These are not well categorized using the Expanded Disability Status Scale (EDSS), the most common measurement of disease severity in MS.^8^ Vision impairment due to factors other than MS progression may confound assessments of vision impairment caused by MS, especially when not evaluated by clinicians familiar with the ophthalmological manifestations of MS.

Limited research has been conducted evaluating the factors that may predict MS-related vision impairment alone. Recently, visual function has been identified as a prognostic factor for MS progression. Optical coherence tomography (OCT) and visual evoked potentials (VEP) show potential for identifying patients with progressive MS, even before reduced visual acuity is evident to the patient.^9,10^ Optical evaluations also show utility in diagnosis of MS.^11^ However, these tools are limited to evaluation after measurable damage has already been done, rather than identification of patients likely to have vision difficulties. A model that predicts patients likely to undergo vision impairment due to MS would provide the opportunity for increased surveillance and early intervention prior to vision impairment.

Our goals for this work are twofold. First, we describe the characteristics of patients with multiple sclerosis who have vision impairment prior to and following MS diagnosis. Second, we develop a model that identifies patients likely to have a diagnosis of vision impairment following diagnosis of MS. To achieve these goals, we utilize Epic Cosmos, a de-identified dataset of electronic health record (EHR) data from 213+ million patients, to train machine learning models, specifically logistic regression (LR) and gradient boosting (LGBM) models, as well as deep learning recurrent neural network (RNN) models to predict patients with MS likely to suffer future vision impairment.

### Background

Vision impairment is recognized as a frequent comorbidity of MS. Vision impairment is not readily captured in existing disease severity scales, potentially leading to underestimation of the number of patients with visual complaints.^12^ Despite this, patients with MS report vision impairment as frequently debilitating, with degree of impairment correlated with the patient’s disease severity.^13^ Existing literature using machine learning to predict outcomes in patients with MS has mainly focused on disease severity and exacerbation. Pinto et al. developed machine learning models to predict risk of MS exacerbation or relapse during the 10 years following the initial diagnosis of MS, achieving AUROC > 80% in some cases.^14^ However, their analysis included features commonly not available within de-identified structured EHR datasets, such as the EDSS and functional sub-scores. Other work focused on predicting vision impairment using genetic markers^15,16^ and imaging data.^17,18^ EHRs are widely available, and the rapid development of predictive models is facilitated by the use of structured data collected and readily available within the EHR. This can also promote efficient adoption of the developed model. However, few studies demonstrated accurate prediction of MS diagnosis or symptom progression using only structured EHR data.

### Significance

Limited research has been done regarding prediction of patients with MS who may be likely to experience vision impairment following their diagnosis of MS. Previous studies have principally focused on prognostic implications using various diagnostic and imaging modalities, not structured clinical data which may indicate the risk of future vision impairment.^11,19^ Recent developments in predictive modeling using RNNs has demonstrated the feasibility of using EHR data to identify patients at risk for adverse health outcomes^20^. Until this point, only López-Dorado et al. have utilized neural networks in predictive modeling for MS. Their 2021 study used optical coherence tomography data to predict diagnosis of MS via a convolutional neural network, achieving 100% accuracy in identifying patients with early MS. However, this work used imaging data alone, and was not focused on disease progression. In this study, we use LR, LGBM, and RNN models trained and validated on the complete medical records of approximately 377,000 patients to identify patients at risk of vision impairment diagnosis following diagnosis of MS. The 377,000-patient population was drawn from a 213+ million patient data set of the United States population, facilitating extensibility of the model to clinical support tools.

### Implications of All the Available Evidence

Vision impairment is a significant problem for patients diagnosed with MS. Prediction of patients likely to be diagnosed with vision impairment can facilitate closer monitoring of patients, selection of disease modifying therapies, and provide insight into how the disease course may affect visual function.^21^ Achieving AUROC of ∼80%, this model demonstrates the feasibility of EHR-based clinical decision support to help physicians identify patients at risk for vision impairment.

## MATERIALS AND METHODS

### Cohort Development

We first evaluated common physical presentations experienced by patients with multiple sclerosis. This was accomplished by identifying patients with multiple sclerosis who also had diagnoses included in the EDSS, such as paraplegia, sensory impairment, incontinence, and vision impairment. Vision impairment was the most common presentation found in this preliminary analysis (Table 1), and we selected it as our main disease of interest in this study.

**Table 1:** Number of patients with physical presentations associated with MS.

| Diagnosis | Number of Patients | % of Total<br>MS *n*= 450,450 |
| --- | --- | --- |
| **Vision Impairment (including optic neuritis)** | 77,001 | 17.09% |
| *Vision loss without optic neuritis* | 56,612 | 12.57% |
| *Optic Neuritis* | 20,389 | 4.53% |
| **Urinary Incontinence** | 68,411 | 15.19% |
| **Vision loss without optic neuritis** | 56,612 | 12.57% |
| **Hemiplegia / Hemiparesis** | 26,098 | 5.79% |
| **Dementia** | 18,607 | 4.13% |
| **Quadriplegia** | 8,516 | 1.89% |

Diagnoses within Cosmos are identified using Cosmos-specific diagnosis codes, primarily based on Intelligent Medical Object terminologies.^22^ These diagnosis codes are linked to other standard diagnosis terminologies, such as SNOMED and ICD-10, and diagnosis code sets, which group related diagnoses. Diagnosis code sets released by Epic were used for preliminary grouping of disease presentations (*Table 1*). The diagnoses included in the vision loss and optic neuritis code sets underwent review by a neurologist for clinical relevance to MS and were divided into diagnoses definitively related to neurological disease and those related to non-neurological or ocular-motor vision impairment (*Supplemental Material A*).

All analyses were completed using the 8/30/2023 Cosmos Expert Determined De-Identified Dataset. The DiagnosisEventFact table was first queried for patients with a diagnosis contained within the Cosmos diagnosis code set for Multiple Sclerosis, which has diagnoses listed in SNOMED concept 24700007, “Multiple sclerosis (disorder)”. No date limit was set. This identified 450,450 patients. We then identified patients from the MS cohort who also had a diagnosis of MS-related vision loss. 70,592 patients had at least one diagnosis from the vision impairment code sets in addition to multiple sclerosis. 366,422 patients had multiple sclerosis and no vision loss diagnosis.

Our goal was to identify patients with multiple sclerosis likely to have a vision loss event occur following their initial diagnosis. This made patients who had a vision loss diagnosis *prior* to multiple sclerosis diagnosis unsuitable for model training. 28,311 patients with a vision loss or optic neuritis diagnosis had their first diagnosis occur prior to multiple sclerosis diagnosis. This is consistent with the known tendency of optic neuritis to occur prior to or concurrently with diagnosis of multiple sclerosis. These patients were separated from the training dataset, since their previous diagnosis could bias the model, and were used only for model evaluation.

Data validity checks were performed on all identified patients. Patients were excluded for a variety of reasons. These included if there was not at least one visit prior to the outcome date of for the patient. Outcome date was designated as the date of vision loss diagnosis (cases) or the most recent encounter (controls). Patients who had a non-MS related vision loss diagnosis *and* a later definitively MS-related vision loss diagnosis were included as cases; however, their outcome date was set to be the visit prior to the non-MS-related vision loss diagnosis to avoid model overfitting of the non-MS-related diagnoses. All exclusion criteria are detailed in *Figure 1*.

**Figure 1:**
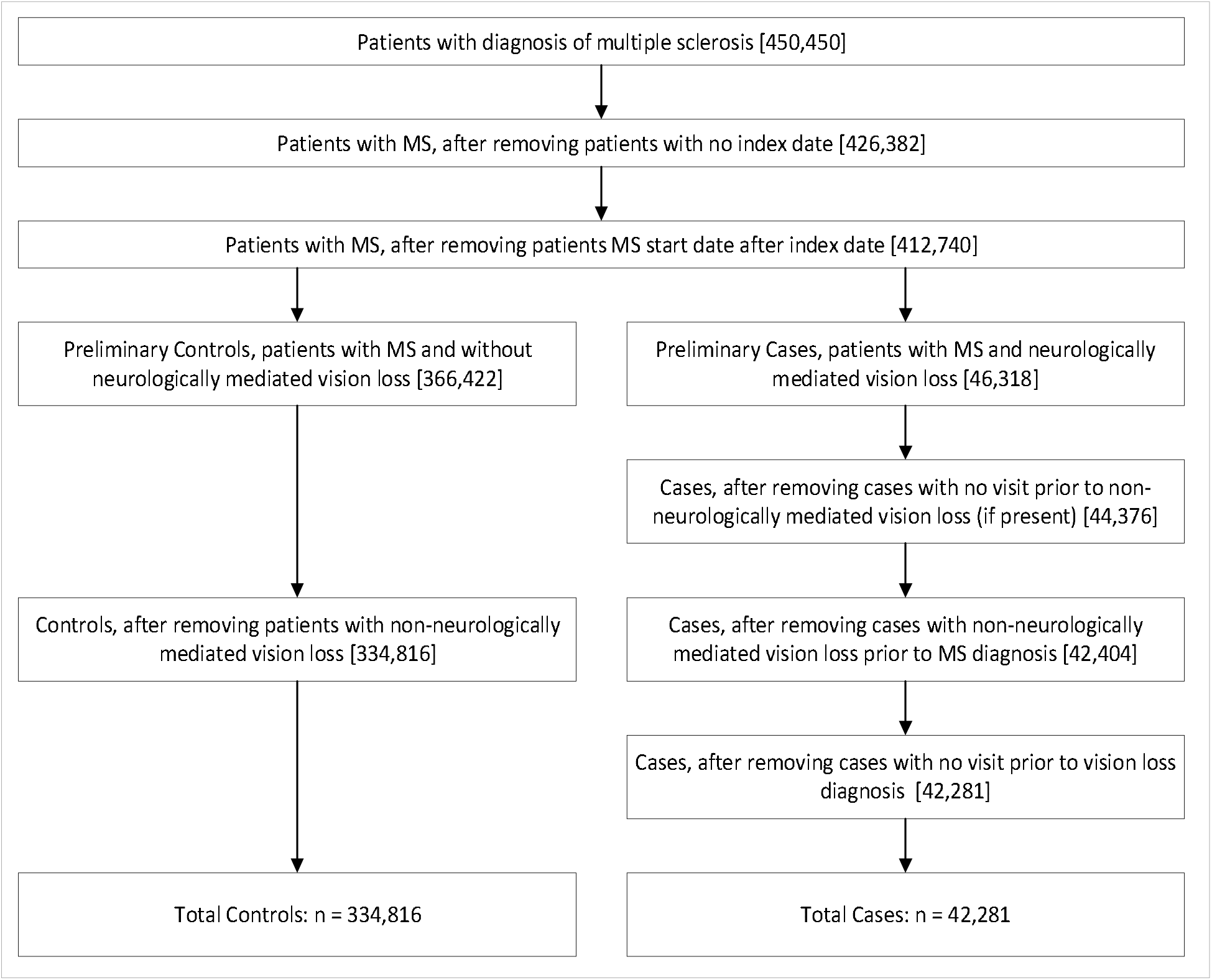
Patient exclusion criteria & cohort determination.

This provided 42,281 patients who had first time vision loss diagnosis following diagnosis of multiple sclerosis (cases) and 334,816 patients with multiple sclerosis and no vision loss diagnosis (controls).

### Data preparation

Cosmos contains an expansive variety of EHR-originated data. This includes discrete features like demographics, medications ordered, diagnoses, laboratory results, and procedures performed. Data that is difficult to de-identify, such as radiology reports or clinical notes, are not available in Cosmos. For this study the lack of discriminatory radiological assessments and direct functional assessment of patient disability limited the ability to calculate commonly used measures of MS severity, such as the EDSS. This led to the use of other features available in Cosmos to develop the model. All diagnosis events, medication orders, and relevant demographic data were included in the model’s dataset. Any value marked as deleted or that otherwise failed data validity checks was not included.

Diagnoses of all types, including Problem List and Encounter diagnoses, were included. In cases where a single diagnosis code mapped to multiple ICD-10 codes, all ICD-10 values were included.

Medications were drawn from inpatient medication orders, clinic medication administrations, and outpatient prescriptions. Medications were limited to the simple generic name of the relevant medication and the date on which it was ordered or prescribed. Dose, frequency, and route were not considered. However, any new medication was included on the order date, allowing for monitoring of disease therapy over time and the introduction of various disease modulating therapies.

Demographic features with patient distributions are listed in *Table 2*. The socioeconomic social vulnerability index is a measure of socioeconomic vulnerability drawn from the published value of the patient’s zip-code of residence^23^. Higher values indicate greater socioeconomic vulnerability.

**Table 2:** Demographic Features.

| **Feature** | **Control** | **Case** |
| --- | --- | --- |
| **Number of patients**<br>**(% of total)** | 334,816 (88.79%) | 42,281 (12.54%) |
| **Sex (*n*, % of total)** |  |  |
| Female | 249,345 (74.47%) | 33,073 (78.22%) |
| Male | 85,266 (25.47%) | 9,203 (21.77%) |
| Other/Unknown | 205 (0.06%) | 5 (0.01%) |
| **Race (*n*, % of total)** |  |  |
| White | 263,653 (78.75%) | 32,418 (76.67%) |
| Black or African<br>American | 44,321 (13.24%) | 7,505 (17.75%) |
| American Indian or<br>Alaska Native | 2,278 (0.68%) | 422 (1.00%) |
| Asian | 2,984 (0.89%) | 366 (0.87%) |
| Native Hawaiian or<br>Other Pacific Islander | 456 (0.14%) | 77 (0.18%) |
| Other/Unknown | 21,124 (6.31%) | 1,493 (3.53%) |
| **Socioeconomic SVI**<br>**(Weighted Average)** | 0.4415 | 0.4621 |

### Models

Source code for model development was taken from the publicly available Pytorch_EHR pipeline used in our earlier CovRNN study.^20^ This source code underwent minor modifications to conform with Cosmos standards and the goals of this study. The core model used for prediction was based on a bi-directional gated-recurrent unit (Bi-GRU) neural network. This model offers advantages for clinical prediction dependent on temporality of events, particularly when using EHR-derived data.^24^ Python 3.10.8 was used for all analyses, with the following packages: Pandas 1.5.3, NumPy 1.24.3, LightGBM 3.3.5, Scikit-learn 1.2.2, Scipy 1.10.1, Statsmodels 0.14.0, and Torch 2.0.1.

For prediction of the binary event of vision impairment diagnosis, the RNN was compared to light gradient boosting machine and logistic regression. All analyses of Cosmos data was completed using a 2.7GHz virtual machine with 32GB of RAM. GPU usage was not available within the Cosmos enclave. This limited the computational and memory capabilities of our implementation, especially with our decisions to not utilize data parallelism and to keep the same features level of granularity for all models. As a result, the training set for the non-RNN models was limited to the top 15,000 most frequent features and 100,000 randomized patients, approximately 1/4^th^ of the total 377,097 patient population.

Three versions of the model were trained for three specific clinical tasks. The “MS-First Diagnosis Model” trained on patient data prior to MS diagnosis. The second “MS-Progress Model” trained on the full patient trajectory until the visit prior to VI diagnosis (cases) or visit prior the most recent encounter (controls). The actual encounter with VI diagnosis was not included in the dataset to prevent the VI diagnosis from being used as a feature. The third “MS-General Model” was trained on both training datasets used for the MS-First Diagnosis and MS-Progress models. This allowed the model to train on both the entirety of the patients’ record *and* their trajectory until MS diagnosis. The datasets were split into training, testing, and validation sets with 70/20/10 percentages of the total allotted for each group. The same patient split was maintained for all models, ensuring no spillover of testing data into the training or validation sets. All model outcome measurements are reported using the 20% testing set.

We then trained an RNN-based survival model. Since the goal of the survival model was to predict likelihood of vision impairment over time, only the MS-First Diagnosis data was used to train the model. Time to event (TTE) was defined as the number of days between MS diagnosis and VI diagnosis (cases) or the most recent encounter in Cosmos (controls). Patients were divided into high (71 – 100^th^ percentile), medium (31-70^th^ percentile), and low (0 – 30^th^ percentile) risk cohorts based on the predicted survival score for Kaplan Kaplan-Meier curve visualization.

All models were evaluated to determine the most important features which contributed to the identification of patients as cases. For logistic regression, beta coefficients were used to assess for possible label leakage and ensure training set validity. The LGBM model was similarly evaluated using lightgbm package’s “plot importance” feature. The RNN model was evaluated using the integrated gradient approach, which does not require gradient back-propagation and provides for extensibility between models.^25^

Each binary and survival prediction model was tested using the patient longitudinal data until their first MS diagnosis encounter (MS-First Diagnosis) and the full patient longitudinal trajectory until the visit preceding the outcome or patient censoring (MS-Progress). The train/test/validation split was maintained with the same patients between the different timepoints, ensuring that training data was not tested in either version of the model.

Model performance was evaluated using area under the receiver operator curve (AUROC), area under the precision recall curve (AUPRC), Brier score for model calibration, and F1-score at a threshold of 10%. The survival model was evaluated using the concordance index. For the precision recall curve, the overall prevalence of vision loss diagnosis in the training set was 11.21%. Due to exclusion of patients with no data prior to MS diagnosis in the MS-First Diagnosis cohort, the MS-First Diagnosis testing set had a slightly different prevalence of vision loss diagnosis at 11.08%.

The binary outcome models were also evaluated using a held-out population of patients with VI diagnosed prior to MS diagnosis. This group is particularly relevant since VI is frequently the first symptom of MS, leading to a VI diagnosis prior to MS diagnosis. All patients in this cohort were later diagnosed with MS. All data prior to the MS-related VI diagnosis or, if present, non-MS related VI diagnosis, were included in the testing set. Since all patients were diagnosed with VI, AUROC could not be calculated. Instead, sensitivity was determined, calculated as the number true positives identified divided by the number of true positives and false negatives, at a threshold of 10%.

Determination of non-human subject research was obtained from the Western Michigan University Homer Stryker M.D. School of Medicine IRB.

## RESULTS

*Table 3* and *Table 4* display basic model performance measures for the MS-First Diagnosis, MS-Progress, and MS-General trained models. Broadly, the models trained on the MS-First Diagnosis dataset performed poorly. The LGBM and RNN performed equivocally, with AUROC, AUPRC, and F-1 metrics within 1% for both models (*Sup. Table 1, Sup. Fig. 1, 2)*. Only the RNN’s Brier score was significantly less than the LGBM, suggesting somewhat better calibration for that model (*Sup.* Fig. 3*, 6*). Overall, the MS-First Diagnosis models did not surpass 68% AUROC when evaluated on the MS-Progress test sets (*Sup.* Fig. 4).

**Table 3:** Models’ performance on binary prediction tasks (AUROC)

| *Table 3: Models' performance on binary prediction tasks (AUROC)* |  |  |  |  |  |  |
| --- | --- | --- | --- | --- | --- | --- |
|  | Prediction at first MS diagnosis |  |  | Prediction at most recent visit |  |  |
|  | LR | LGBM | RNN | LR | LGBM | RNN |
| MS-First Diagnosis | 0.591 | 0.621 | 0.632 | 0.649 | 0.681 | 0.679 |
| MS-Progress | 0.607 | 0.628 | 0.627 | 0.686 | 0.728 | **0.795** |
| MS-General | 0.604 | 0.628 | **0.644** | 0.677 | 0.720 | 0.788 |

**Table 4:** Binary and Survival models’ performance on prediction tasks (C-index)

|  |  |  |
| --- | --- | --- |
| Model | Prediction at first MS diagnosis | Prediction at most recent visit |
| MS-First Diagnosis | **0.63** | 0.581 |
| MS-Progress | 0.575 | **0.75** |
| MS-General | 0.613 | 0.729 |

*Supplementary Table 2* displays performance measures for the models trained on the entirety of the patient trajectory until the visit preceding the outcome (MS-Progress). The RNN based models exhibited superior performance in all measures, achieving AUROC of 79.6% (*Sup.* Fig. 4). The RNN also had a significantly better AUPRC than both models at 40.5%, 12% greater than the LGBM (*Sup.* Fig. 5). The MS-Progress models performed worse on the MS-First Diagnosis dataset than the models trained on the MS-First Diagnosis data alone, suggesting the patient’s EHR-documented course following diagnosis progresses substantially differently than that prior to diagnosis. Calibration curves were again poorly aligned when tested on MS-First Diagnosis data, but the RNN exhibited excellent calibration when tested on the full patient trajectory (*Sup.* Fig. 3, *6*).

The MS-General model (*Sup. Table* 3) demonstrated somewhat balanced performance across both testing datasets. The RNN performed better in all measures compared to the LR and LGBM models but did not outperform the MS-Progress RNN. Performance at the MS diagnosis remained relatively poor with an AUROC of 64.4%, though with a ∼1.1% AUROC gain over the MS-First Diagnosis trained model.

The best performing survival model was the MS-Progress model when evaluated during the patient’s most recent visit with a concordance index (c-index) of 75%, while the MS-First Diagnosis survival model showed a c-index of 63% when used at the initial MS diagnosis. Despite this, discrimination for patients who were at high, medium, or low risk for vision impairment at 5 years was strong, as illustrated in the Kaplan-Meier survival curve (*Sup.* Fig. 7). This suggests that the ability for broad discrimination is still present within the model, and that the model may perform well in tasks not requiring precise diagnostic accuracy.

We additionally evaluated the held-out group of patients diagnosed with MS-related VI prior to diagnosis of MS. *Table 5* details sensitivity of the models at 10% threshold. The MS-General model achieved the best sensitivity across all ML model types; however, high sensitivity may come at the cost of discriminative accuracy when patients without VI are evaluated.

**Table 5:**
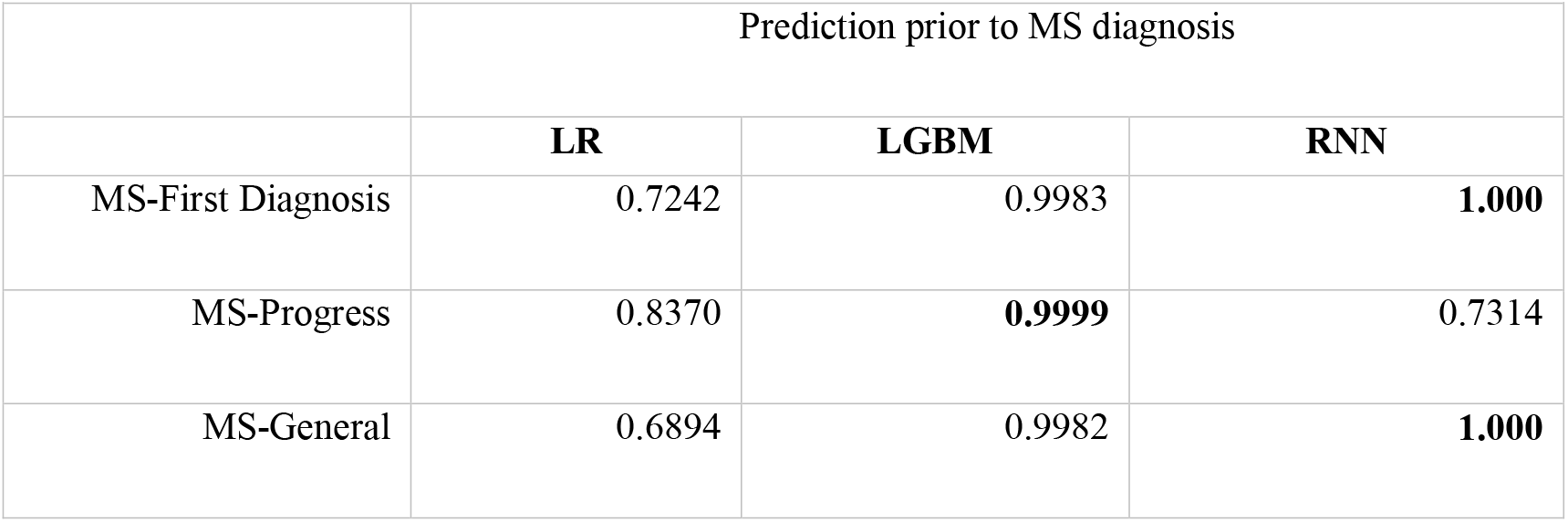
Sensitivity of models predicting VI prior to MS diagnosis at 10% threshold.

|  |  |  |  |
| --- | --- | --- | --- |
|  | Prediction prior to MS diagnosis |  |  |
|  | **LR** | **LGBM** | **RNN** |
| MS-First Diagnosis | 0.7242 | 0.9983 | **1.000** |
| MS-Progress | 0.8370 | **0.9999** | 0.7314 |
| MS-General | 0.6894 | 0.9982 | **1.000** |

## DISCUSSION

Performance of the RNN trained on the entirety of the dataset reached ∼80% AUROC, 5% greater than the LGBM. RNN has shown promise for modeling using large clinical datasets, and demonstrated superiority in this analysis.^20^ The RNN additionally achieved near-perfect model calibration and AUPRC 10% greater than the LGBM, approximately 30% greater than the base case prevalence. However, F-1 of the model only reached ∼40%, which was expected given the known class imbalance of the dataset.

Model performance was strong when considering the entirety of a patient’s medical record. Cosmos provides a detailed view of the discrete features contained in a patient’s record, and any models developed from it are most suited to tools where detailed electronic health record data will provide the majority of data informing predictions. It is clear that predicting future vision impairment at the time of MS diagnosis is a difficult task, and that vision impairment prediction is far more accurate when the chronology of a patient’s MS course is considered.

Notably, our model excluded patients diagnosed with vision loss prior to MS diagnosis. Since ON is frequently the presenting symptom of MS this led to an excluded cohort of patients only ∼1% smaller than the case cohort. These patients are likely to have recurrence of ON at some point following their diagnosis, and present a unique group which should be examined in future model development. This may account for some of the poor performance in models trained using only patients without vision loss prior to diagnosis. Future efforts could include analysis of the time between ON or vision loss events, and the likelihood of vision impairment recurrence following diagnosis.

Cosmos lacks multiple features that provide significant insight into the clinical status of a patient with MS. These include the EDSS, commonly used to measure disease severity, radiological reports, and clinical notes. Existing models have typically utilized at least the EDSS as a measure of disease severity, along with other imaging data from a variety of modalities^19^. The EDSS is made up of a variety of subscores which are potentially captured through diagnosis codes found in Cosmos, but duration and severity of disability due to those conditions is difficult to ascertain. The use of RNN modeling potentially helps improve performance in this regard, since clusters of related symptoms are evaluated more generally than in logistic regression or LGBM models.

A major goal of model development using massive clinical datasets like Cosmos is generalizability of models to other data sources. This model is well suited for deployment within EHR’s for in-workflow clinical decision support. However, it demonstrates difficulty integrating with other data sources. Examination of the features included in a sample registry dataset revealed that the Cosmos level of granularity is not shared by extant disease registries, and that lack of commonly used measures of disease progression is a major consideration for future implementation of the model. As a result of this study, discussions of the feasibility of the inclusion of measures like the EDSS are being undertaken with Cosmos developers. Additionally, further work by our team is planned to harmonize features in Cosmos with those commonly included in the global iMed MSBase MS registry to facilitate model transferability and evaluation on international data.^26^

The ultimate goal of any prognostic model is clinical utility. The clinical utility of identification of patients likely to have VI following diagnosis of MS is directly related to the intervention that can take place based on that knowledge. In the case of MS, this may include more intensive monitoring for vision changes or more aggressive therapy for patients at high risk. While this study approached VI as an isolated outcome, it is likely frequently part of broader MS progression. Further research needs to be done to identify proxies for MS progression that can be derived from EHR data, allowing for a more general model of MS from pre-diagnosis to death to be developed.

## CONCLUSION

RNN models trained on data from Epic Cosmos provide a viable approach for predicting vision impairment in patients with MS. Though previous studies have attempted to facilitate the early identification and diagnosis of patients likely to have vision loss using imaging data, this is the first model to use EHR data to achieve that goal. EHR data is ubiquitous and provides a gestalt view of the patient’s condition chronologically and independent of care specifically related to MS diagnosis. This made it a strong data source for model development, as well as future deployment of this and similar models. Models trained on EHR data are technically suited for deployment within EHR’s, especially those with a data structure and granularity similar to their training dataset. Our use of the entire patient record included in Cosmos minimizes the need for aggressive data cleaning, and use of a massive international population helps avoid the disadvantages of using single center registries for extensible model training.

Our work identified over 377,000 patients, representative of the population of the United States with MS, and trained three machine learning models to predict vision loss. The RNN based models performed best, achieving discriminative accuracy of 79.5% AUROC at time of prediction and 63.1% AUROC at time of diagnosis. LGBM also performed comparably with the RNN, demonstrating feasibility of traditional machine learning approaches for this problem.

While the RNN achieved ∼80% AUC when trained and tested on the entirety of the dataset, there is clearly work remaining. The most clinically useful point for predicting future vision loss is at the diagnosis of MS, not immediately prior to the occurrence and diagnosis of some form of vision impairment. All models developed struggled with the limited MS-First Diagnosis dataset, and it is unclear if different training approaches may improve performance. Additionally, the RNN does not lend itself to important feature selection, making it difficult to operationally implement without multiple iterations of training on less granular data.

Future work with this model will focus on enhancing the ability for disparate datasets to be used for both testing and patient evaluation. Since the patient’s clinical course, and therefore medical record, is a highly heterogenous data source, the extent and complexity of the record may influence the model’s predictions in unidentified ways. Evaluating the model using different, more limited datasets will help validate the model and clinical relevance in cases where a provider does not have extensive historical records of a patient’s treatment. Overall, RNN’s have demonstrated potential for predicting clinical events and this study supports their utility in treating patients with MS.

## Data Availability

data used in this study came from Epic Cosmos, a community collaboration of health systems representing over 220 Million patient records from over 1277 hospitals and 27200 clinics. The current count values for patients, hospitals, and clinics are available on cosmos.epic.com. All data produced in the present work are contained in the manuscript.

https://cosmos.epic.com/

## Acknowledgements

We thank the entire Epic Cosmos team, especially Laura Hosmer, Harry Freedman, and Meghan Howat for their contribution to this work and for their dedication to making research using Cosmos possible.

## Supplementary Tables

**Supplementary Table 1:** Model Performance – MS-First Diagnosis Model.

| Model | Logistic Regression |  | LGBM |  | RNN |  |
| --- | --- | --- | --- | --- | --- | --- |
| Metric | Prediction at first MS diagnosis | Prediction at most recent visit | Prediction at first MS diagnosis | Prediction at most recent visit | Prediction at first MS diagnosis | Prediction at most recent visit |
| AUROC | 0.5907 | 0.6491 | 0.6212 | 0.6805 | 0.6316 | 0.6790 |
| AUPRC | 0.1663 | 0.2296 | 0.1863 | 0.2580 | 0.1851 | 0.2412 |
| F-1 | 0.2446 | 0.2813 | 0.2620 | 0.2984 | 0.2650 | 0.2996 |
| Brier Score | 0.1158 | 0.1127 | 0.2240 | 0.1761 | 0.1064 | 0.1057 |

**Supplementary Table 2:** Model Performance – MS-Progress Model.

| Model | Logistic Regression |  | LGBM |  | RNN |  |
| --- | --- | --- | --- | --- | --- | --- |
| Metric | Prediction at first MS diagnosis | Prediction at most recent visit | Prediction at first MS diagnosis | Prediction at most recent visit | Prediction at first MS diagnosis | Prediction at most recent visit |
| AUROC | 0.6069 | 0.6860 | 0.6276 | 0.7282 | 0.6268 | 0.7956 |
| AUPRC | 0.1524 | 0.2221 | 0.1670 | 0.2807 | 0.1632 | 0.4051 |
| F-1 | 0.2252 | 0.2847 | 0.1986 | 0.2031 | 0.2349 | 0.3697 |
| Brier Score | 0.1054 | 0.0998 | 0.2687 | 0.1984 | 0.1068 | 0.0821 |

**Supplementary Table 3:** Model Performance – MS-General Model.

| Model | Logistic Regression |  | LGBM |  | RNN |  |
| --- | --- | --- | --- | --- | --- | --- |
| Metric | Prediction at first MS diagnosis | Prediction at most recent visit | Prediction at first MS diagnosis | Prediction at most recent visit | Prediction at first MS diagnosis | Prediction at most recent visit |
| AUROC | 0.6041 | 0.6771 | 0.6286 | 0.7200 | **0.6435** | **0.7876** |
| AUPRC | 0.1502 | 0.2139 | 0.1675 | 0.2706 | **0.1793** | **0.3878** |
| F-1 | 0.2289 | 0.2837 | 0.1988 | 0.2038 | **0.2453** | **0.3602** |
| Brier Score | 0.1036 | 0.1009 | 0.2417 | 0.1864 | **0.0958** | **0.0829** |

## Supplementary Figures

**Figure S1:**
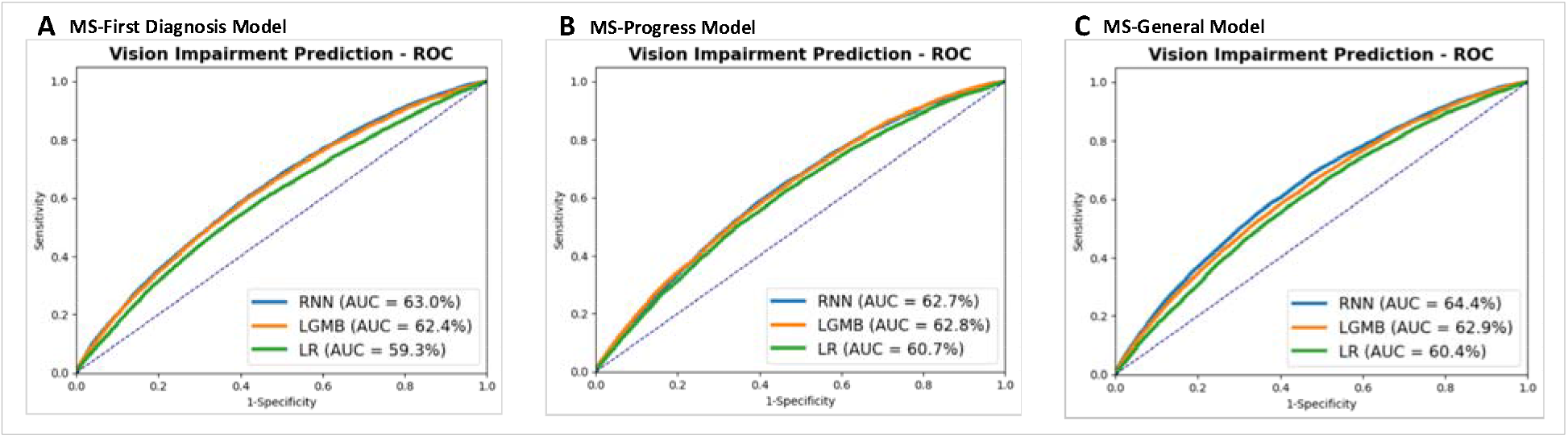
AUROC curve of the MS-Diagnosis model (A), MS-Progress model (B), and MS-General model (C) tested on the MS-First Diagnosis test set.

**Figure S2:**
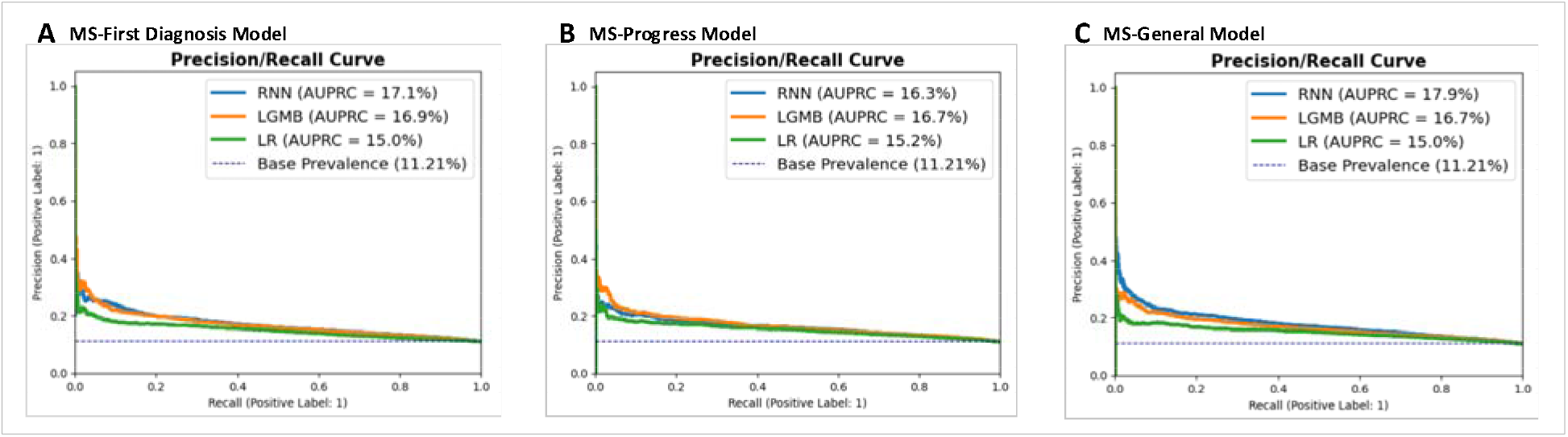
AUPRC curve of the MS-Diagnosis model (A), MS-Progress model (B), and MS-General model (C) tested on the MS-First Diagnosis test set.

**Figure S3:**
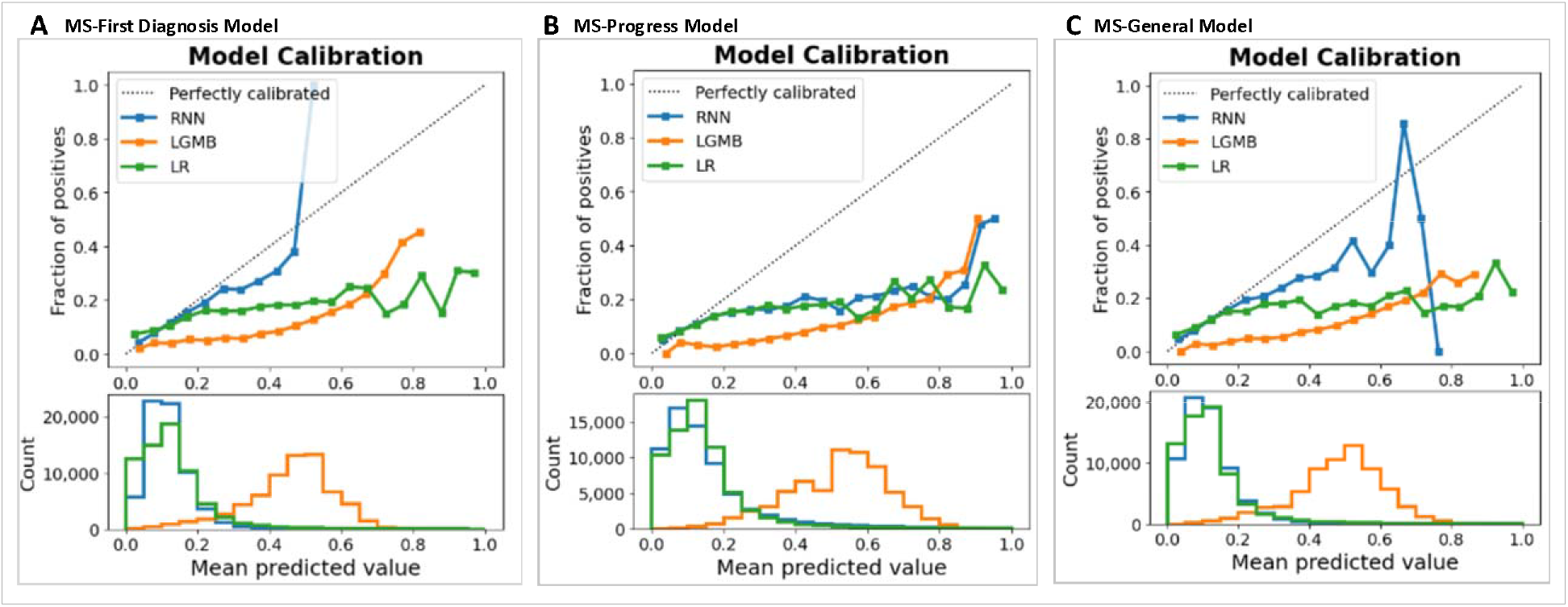
Calibration curve of the MS-Diagnosis model (A), MS-Progress model (B), and MS-General model (C) tested on the MS-First Diagnosis test set.

**Figure S4:**
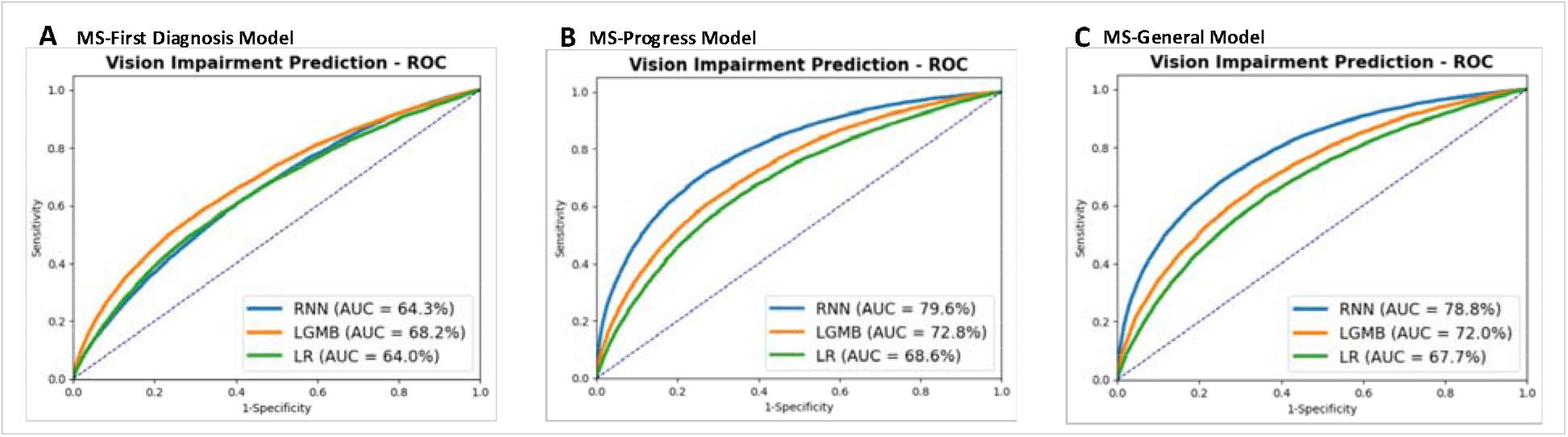
AUROC curve of the MS-Diagnosis model (A), MS-Progress model (B), and MS-General model (C) tested on the MS-Progress test set.

**Figure S5:**
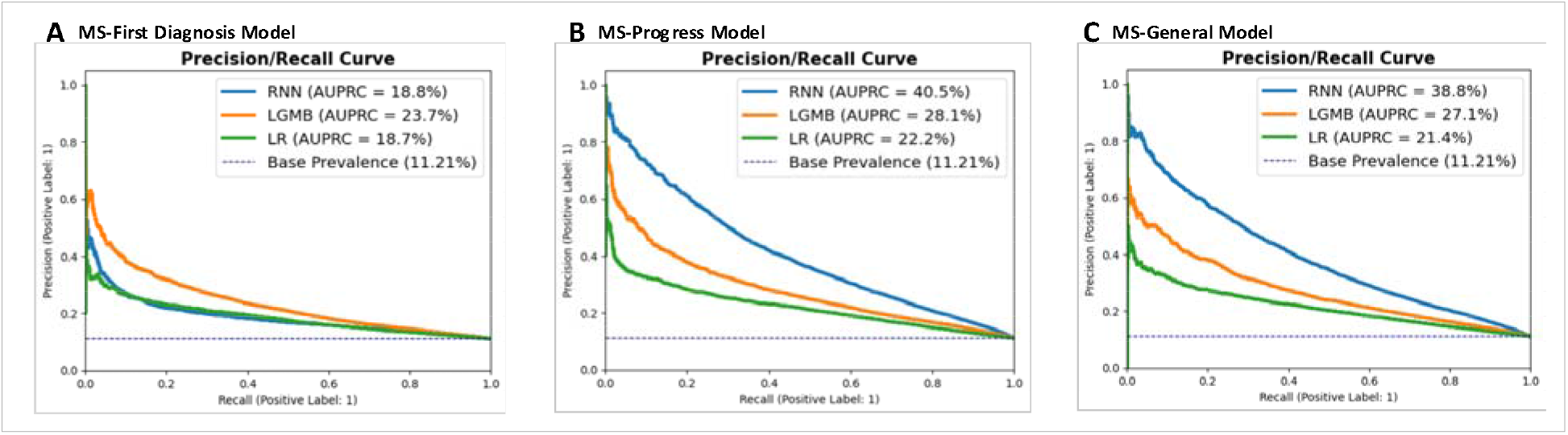
AUPRC curve of the MS-Diagnosis model (A), MS-Progress model (B), and MS-General model (C) tested on the MS-Progress test set.

**Figure S6:**
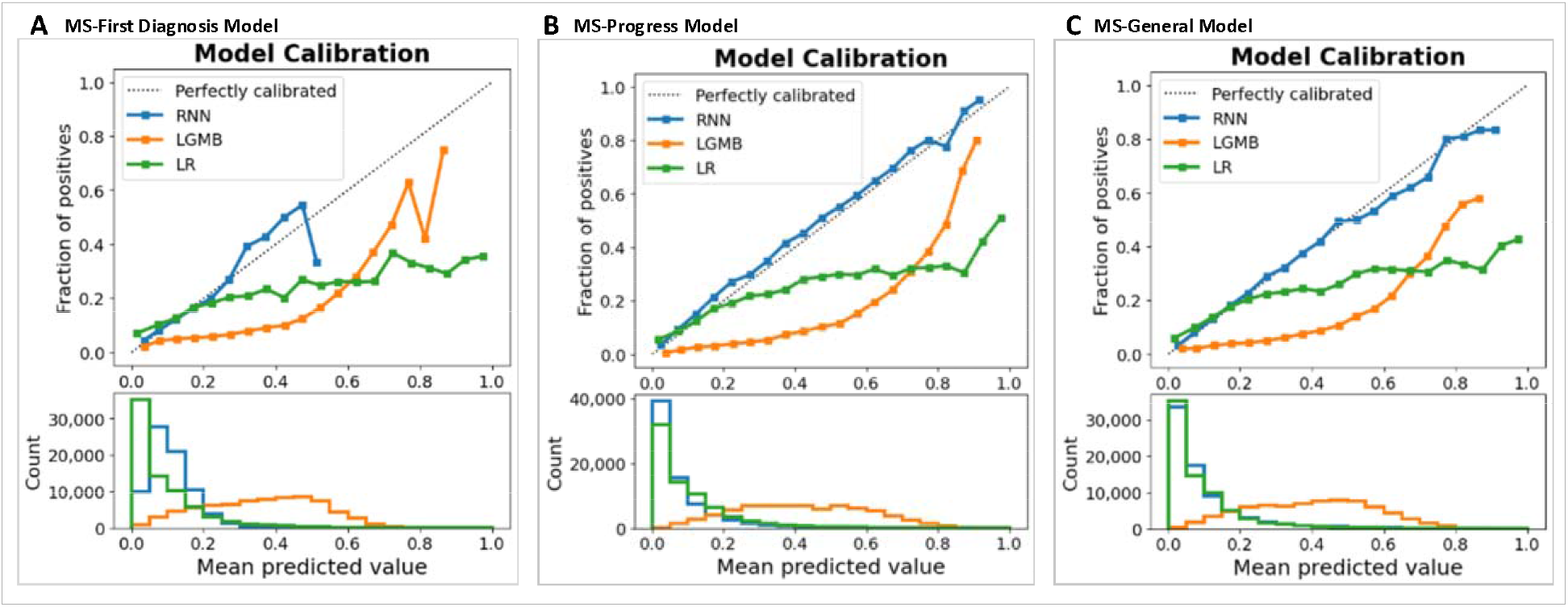
Calibration curve of the MS-Diagnosis model (A), MS-Progress model (B), and MS-General model (C) tested on the MS-Progress test set.

**Figure S7:**
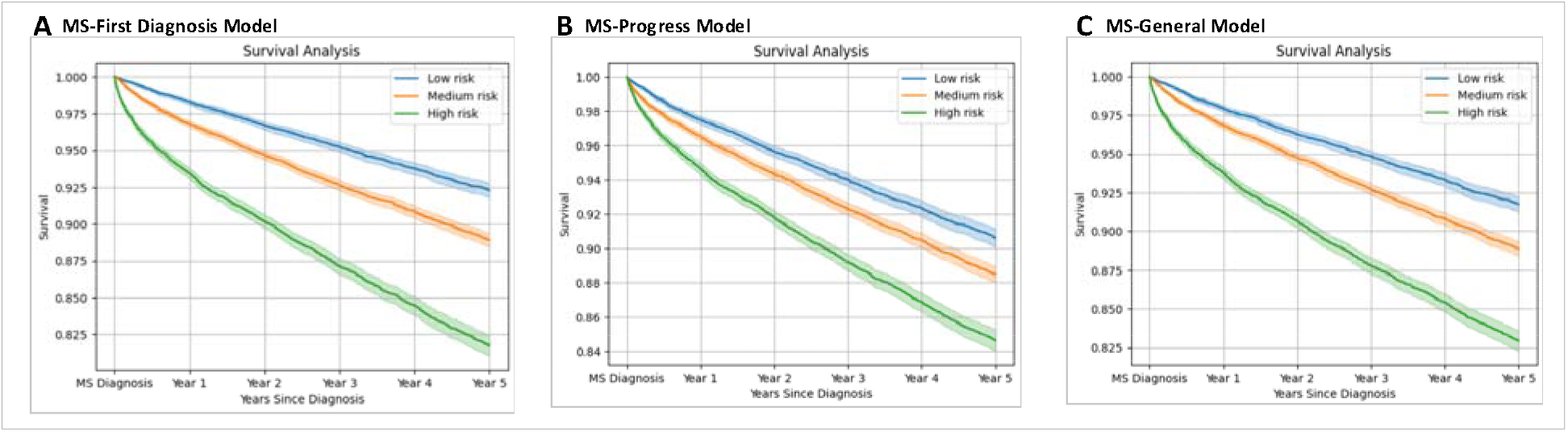
Kaplan-Meier curve of patients likely to have vision impairment within 5 years of the MS-Diagnosis model (A), MS-Progress model (B), and MS-General model (C) tested on the MS-First Diagnosis test set.

